# The epidemiology of knee injuries in New Zealand, 2015–2024

**DOI:** 10.64898/2026.08.27.26361563

**Authors:** Yana Pryymachenko, Ross Wilson, J. Haxby Abbott

**Affiliations:** Centre for Musculoskeletal Outcomes Research, Department of Surgery and Critical Care, University of Otago, Dunedin, New Zealand

## Abstract

**Background:** Little evidence is available on the epidemiology of different knee injuries at a whole-of-population level. The objective of this article is to provide accurate estimates of knee injury incidence by harnessing the unique comprehensive, population-wide data of New Zealand’s universal no-fault injury insurance provider, the Accident Compensation Corporation (ACC).

**Methods:** We obtained insurance claims data from ACC covering all knee injury insurance claims approved between 2015 and 2024. We calculated the number of injuries and the incidence rate per 100 000 population, by injury type, year, sex, ethnicity, and age.

**Results:** The total number of injuries increased from 184 710 (4 067 per 100 000 population) in 2015 to 244 155 (4 701 per 100 000) in 2024. The most common injuries were other/unspecified ligament sprains, contusions, and collateral ligament sprains. Ligament and cartilage injuries were more common for males than for females, while contusions were more common for females. Ligament tears and dislocations were more common in younger people (15 to 35 years of age), while cartilage injuries were more common at older ages (40 to 65 years).

**Discussion and Conclusions:** The rate of knee injuries observed in this study was higher than previously reported in other settings, probably due to broader coverage of injuries treated in primary and community care settings. A broad range of injuries were common, including those that have received less attention in the epidemiological literature to date. More research is needed on the prevention, burden, and outcomes of different knee injuries, beyond a narrow focus on cruciate ligament injuries.

## Background

To establish effective and value-for-money injury prevention and management strategies, it is necessary to first understand the scope and nature of the problem: the rate of occurrence of knee injuries, and their distribution across the population and over the life course. This information can assist in identifying health service delivery needs, assessing the potential impact of injury prevention initiatives, and targeting both research and health service investment where it is most needed. However, there is currently little research on the epidemiology of various types of knee injuries at a whole-of-population level. Existing studies have either relied on data from specific healthcare settings (e.g., emergency department visits or hospital admissions^1,2^) or subnational regions^3^, or focused on certain subpopulations (e.g., children or sports participants^4,5^) or a particular type of knee injury (mostly cruciate ligament injury^6,7^).

New Zealand’s (NZ) comprehensive national injury insurance scheme provides a unique opportunity to address this research gap. The Accident Compensation Act (1972), the first legislation of its type in the world, established a unique social contract by creating a comprehensive, state-run, no-fault personal injury insurance scheme, in exchange for essentially barring common-law personal injury claims for all people injured in New Zealand. The claims database of the Accident Compensation Corporation (ACC), which administers the insurance scheme, thus includes almost all injuries sustained in New Zealand, irrespective of severity, setting, or cause. These data can also be linked with nationwide, individual-level administrative data, allowing detailed demographic analysis of the injuries.

This study seeks to fill the existing gap in the literature by investigating the population-wide frequencies and incidence rates of a range of knee injuries in New Zealand over the period 2015 to 2024, and their distribution across demographic strata.

## Methods

### Data

All data were obtained from the Integrated Data Infrastructure (IDI), a linked research database maintained by Stats NZ, New Zealand’s national statistics agency, containing individual-level data from government administrative datasets, Stats NZ surveys, and non-governmental organisations^8^.

Within the IDI, data on knee injuries were collected from the ACC claims dataset, which covers all injury claims made to the ACC since 1994. As the ACC is the sole provider of injury insurance in New Zealand (covering most injury costs on a no-fault basis), the claims database includes the vast majority of knee injuries sustained in New Zealand, irrespective of severity, setting, or cause.

The age, sex, and ethnicity of claimants were identified from the linked Personal Details dataset in the IDI. Population data were taken from the estimated New Zealand resident population datastanalyses were conductede, and refer to the estimated resident population of New Zealand as at 30 June each year.

### Injury identification

We extracted data on all knee injuries sustained between 2015 and 2024. The claims dataset records diagnoses using three different clinical coding systems: the International Classification of Diseases, 9th revision (ICD-9), the International Classification of Diseases, 10th revision (ICD-10), and Read codes. Knee injuries were identified if they were recorded using any of these systems. We grouped injuries into soft tissue injuries (cruciate ligament injuries, classified as anterior cruciate ligament tears, posterior cruciate ligament tears, or sprains; collateral ligament injuries, classified as medial or lateral collateral ligament tears or sprains; other or unspecified ligament injuries, classified as tears or sprains; and cartilage injuries, primarily meniscus tears), fractures and dislocations, and other knee injuries (classified as contusions, crush injuries, or other or unspecified injuries). The codes used to identify each type of injury are reported in Table A1.

One claim may have multiple diagnoses for different knee injuries, all of which were counted as separate injuries in our analysis. We also calculated the total number of knee injuries, the number of knee injury claims (representing specific injury-causing events), and the number of claims with more than one knee injury.

### Statistical analysis

We calculated the number of injuries and the incidence rate per 100 000 population, for each type of injury, and stratified these by year, sex, ethnicity, and age. Ethnicity was classified as Māori (New Zealand’s indigenous population) and non-Māori. Yearly trends in the number of injuries (total and for each injury type) are presented for the total population, and incidence rates (per 100 000 population) for each sex and ethnicity subgroup. The distribution of knee injuries by age and sex for the entire 10-year study period is presented using population pyramids. Due to the security provisions for data released from the IDI, all counts have been randomly rounded (up or down) to the nearest multiple of three (and rates calculated from these rounded counts).

All analyses were conducted using R (version 4.5.1)^9^.

### Ethics and data availability

This study was approved by the University of Otago Human Research Ethics Committee (Health) [HD20/074]. Access to the anonymised data used in the study was provided by Stats NZ under the security and confidentiality provisions of the Statistics Act 1975; see the full data disclaimer at the end of this article for further details.

The individual-level raw data used in the study are not publicly available due to the strict security provisions of the IDI. Access to the IDI may be made available by Stats NZ to approved researchers. The aggregated data (by injury/year/sex/ethnicity/age strata), and the code used to construct the tables and figures presented here, are available via Zenodo (https://doi.org/10.5281/zenodo.22104854).

## Results

There were 184 710 knee injuries recorded in 2015, increasing to 244 155 in 2024 (Table 1). The most common injuries were other/unspecified ligament sprains (136 437 in 2024), contusions (46 971), and collateral ligament sprains (30 930).

**Table 1:** Annual number of knee injuries in New Zealand, 2015 to 2024.

| Injury | 2015 | 2016 | 2017 | 2018 | 2019 | 2020 | 2021 | 2022 | 2023 | 2024 |
| --- | --- | --- | --- | --- | --- | --- | --- | --- | --- | --- |
| **Soft tissue injuries** |  |  |  |  |  |  |  |  |  |  |
| *Cruciate ligament injuries* |  |  |  |  |  |  |  |  |  |  |
| ACL tear | 3 009 | 3 018 | 3 051 | 3 165 | 3 480 | 2 928 | 2 922 | 2 979 | 3 495 | 4 089 |
| PCL tear | 135 | 126 | 147 | 147 | 177 | 153 | 165 | 180 | 180 | 288 |
| Sprain | 6 195 | 5 859 | 5 550 | 5 733 | 5 661 | 4 917 | 5 118 | 4 860 | 5 121 | 5 352 |
| *Collateral ligament injuries* |  |  |  |  |  |  |  |  |  |  |
| Lateral tear | 117 | 111 | 111 | 108 | 120 | 105 | 120 | 84 | 102 | 123 |
| Medial tear | 357 | 348 | 375 | 360 | 366 | 450 | 453 | 393 | 486 | 696 |
| Sprain | 37 860 | 37 938 | 36 714 | 36 711 | 35 907 | 32 754 | 31 866 | 29 322 | 31 050 | 30 930 |
| *Other/unspecified ligament injuries* |  |  |  |  |  |  |  |  |  |  |
| Tear | 114 | 126 | 111 | 123 | 120 | 159 | 159 | 192 | 189 | 207 |
| Sprain | 72 168 | 79 650 | 84 171 | 88 818 | 95 997 | 94 287 | 99 627 | 105 867 | 120 159 | 136 437 |
| Cartilage injuries | 15 732 | 14 202 | 13 827 | 14 364 | 14 298 | 13 905 | 14 046 | 13 932 | 14 745 | 13 485 |
| **Fractures and dislocations** |  |  |  |  |  |  |  |  |  |  |
| Fracture | 1 530 | 1 599 | 1 611 | 1 560 | 1 467 | 1 326 | 1 209 | 1 098 | 1 131 | 1 053 |
| Dislocation | 1 272 | 1 251 | 1 206 | 1 266 | 1 296 | 1 185 | 1 251 | 1 047 | 1 149 | 1 248 |
| **Other knee injuries** |  |  |  |  |  |  |  |  |  |  |
| Contusion | 43 659 | 44 862 | 45 282 | 46 026 | 46 611 | 42 393 | 43 032 | 41 817 | 45 480 | 46 971 |
| Crush injury | 57 | 69 | 57 | 72 | 81 | 72 | 78 | 72 | 87 | 102 |
| Other/unspecified | 2 502 | 2 652 | 2 799 | 2 931 | 2 808 | 3 144 | 3 366 | 3 822 | 4 470 | 3 180 |
| **Total** | **184 710** | **191 814** | **195 006** | **201 387** | **208 389** | **197 772** | **203 412** | **205 668** | **227 844** | **244 155** |
| Claims | 165 678 | 173 139 | 175 536 | 180 882 | 187 383 | 177 606 | 181 854 | 183 567 | 203 337 | 218 289 |
| Multiple-injury claims | 16 479 | 16 233 | 16 815 | 17 679 | 18 000 | 17 262 | 18 483 | 18 873 | 20 643 | 21 405 |

Abbreviations: ACL = Anterior cruciate ligament, PCL = Posterior cruciate ligament.

The total knee injury incidence rate increased by 16% (from 4 067 to 4 701 per 100 000 population) over the study period (Figure 1 and Table A2). The largest increases occurred for posterior cruciate ligament tears (87%), medial collateral ligament tears (70%), other or unspecified ligament tears (59%) and sprains (65%), and crush injuries (56%). Significant declines in incidence rates were observed for fractures (−40%), collateral ligament sprains (−29%), cartilage injuries (−25%), and cruciate ligament sprains (−24%).

**Figure 1:**
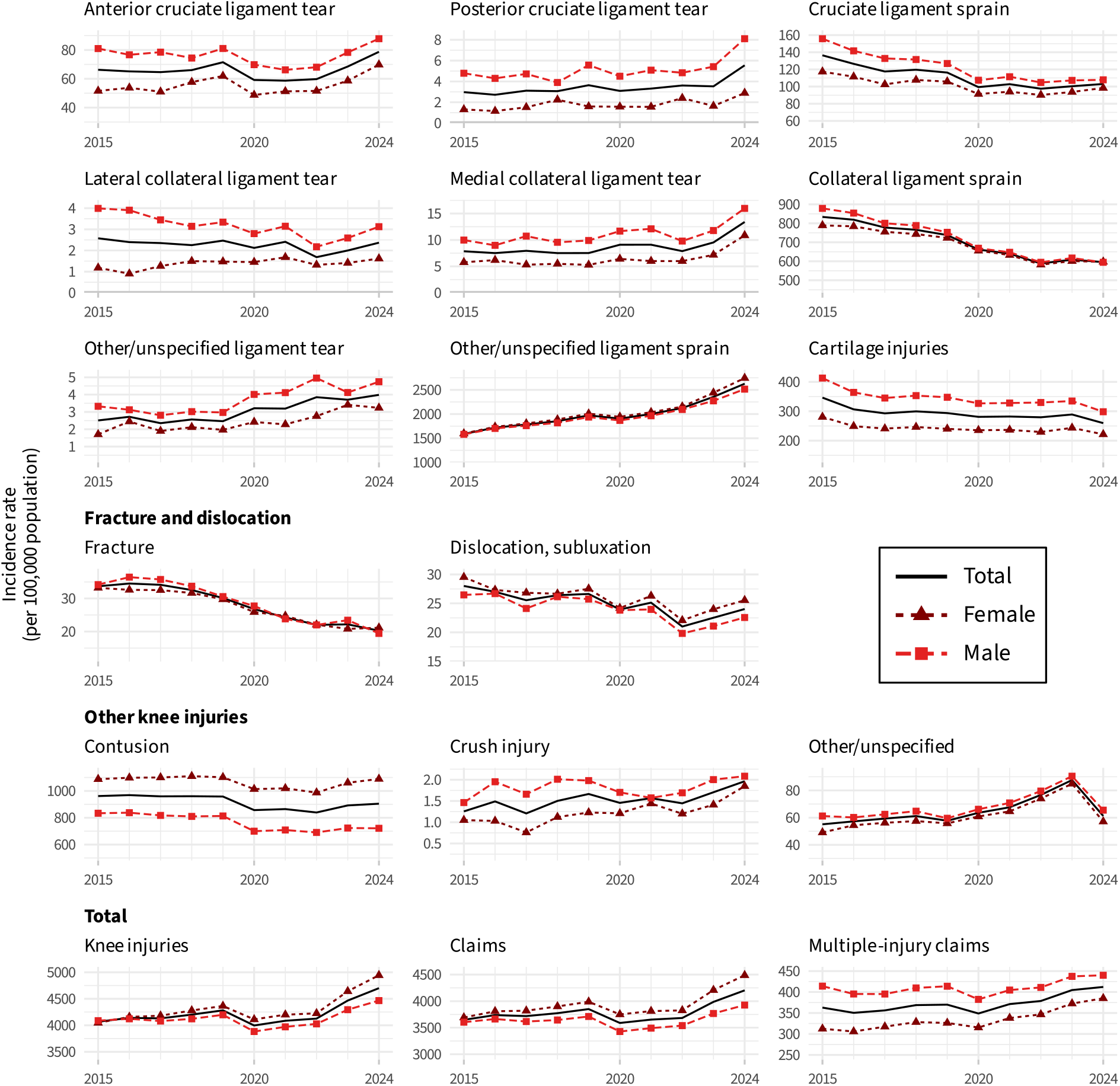
Annual incidence of knee injuries in New Zealand, 2015 to 2024, by sex

Males had higher incidence rates than females for soft tissue injuries, except for other/unspecified ligament sprains, while females had higher incidence rates for contusions and dislocations. The increase in incidence rate (of all knee injuries) over the study period was greater for females (22%; 4 047 to 4 944 per 100 000) than for males (9%; 4 088 to 4 467 per 100 000).

Māori had a higher incidence rate of ligament tears and dislocations than non-Māori, but lower rates of ligament sprains, cartilage injuries, and contusions (Figure 2). The trends over the study period were similar for Māori and non-Māori.

**Figure 2:**
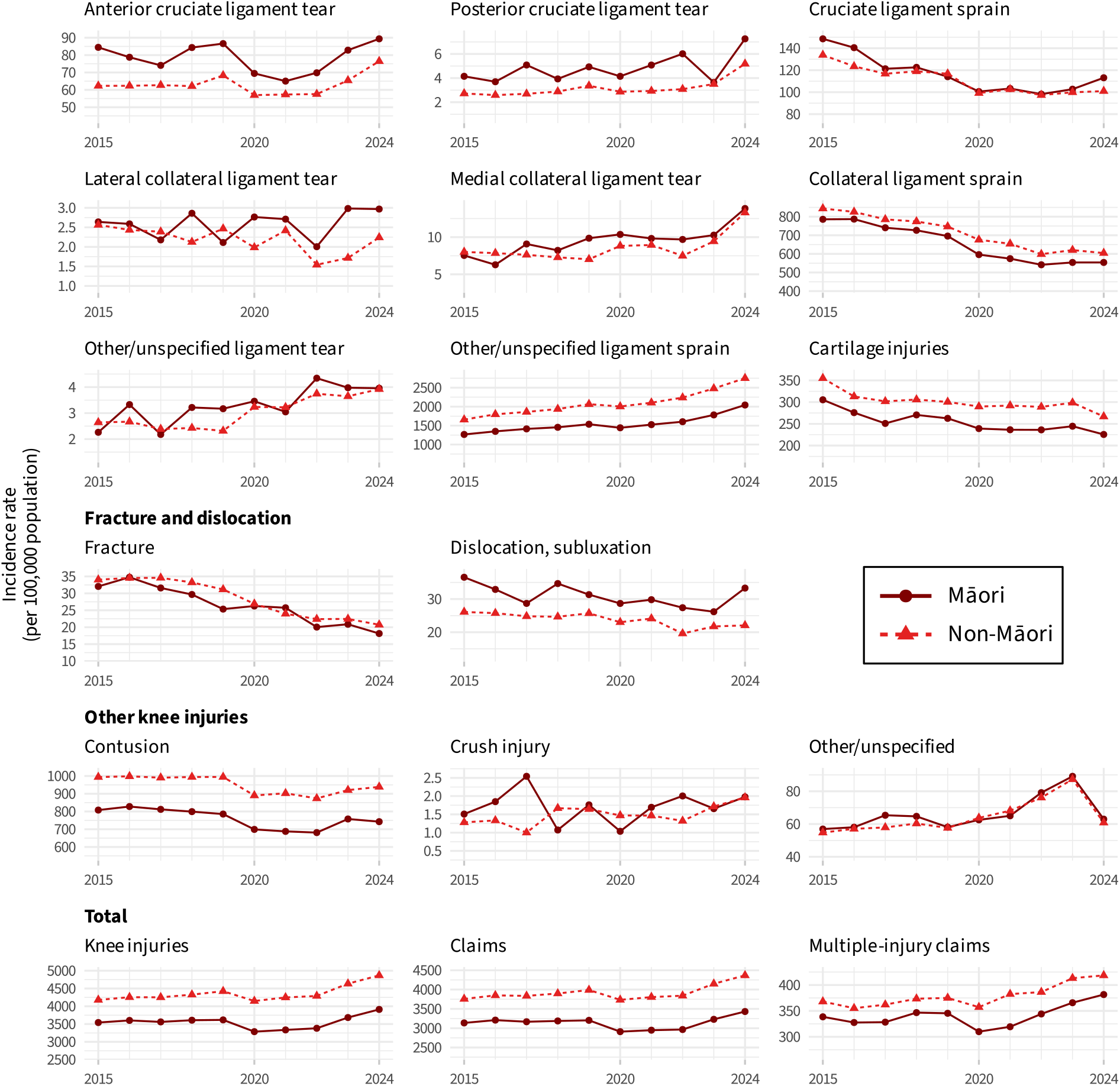
Annual incidence of knee injuries in New Zealand, 2015 to 2024, by ethnicity

The distribution of knee injuries by age and sex varied with knee injury type (Figure 3). Ligament tears and dislocations were more common among people aged 15 to 35, while cartilage injuries were more prevalent at 40 to 65 years of age. Contusions and fractures were more common at age 50 and above for women but not for men. The overall rate of knee injuries was around 4000 per 100 000 population in all age groups 15 years and over, being somewhat higher for men aged 15 to 30 and women aged 50 and over. For children aged under 15, incidence rates were low for all injuries except fractures.

**Figure 3:**
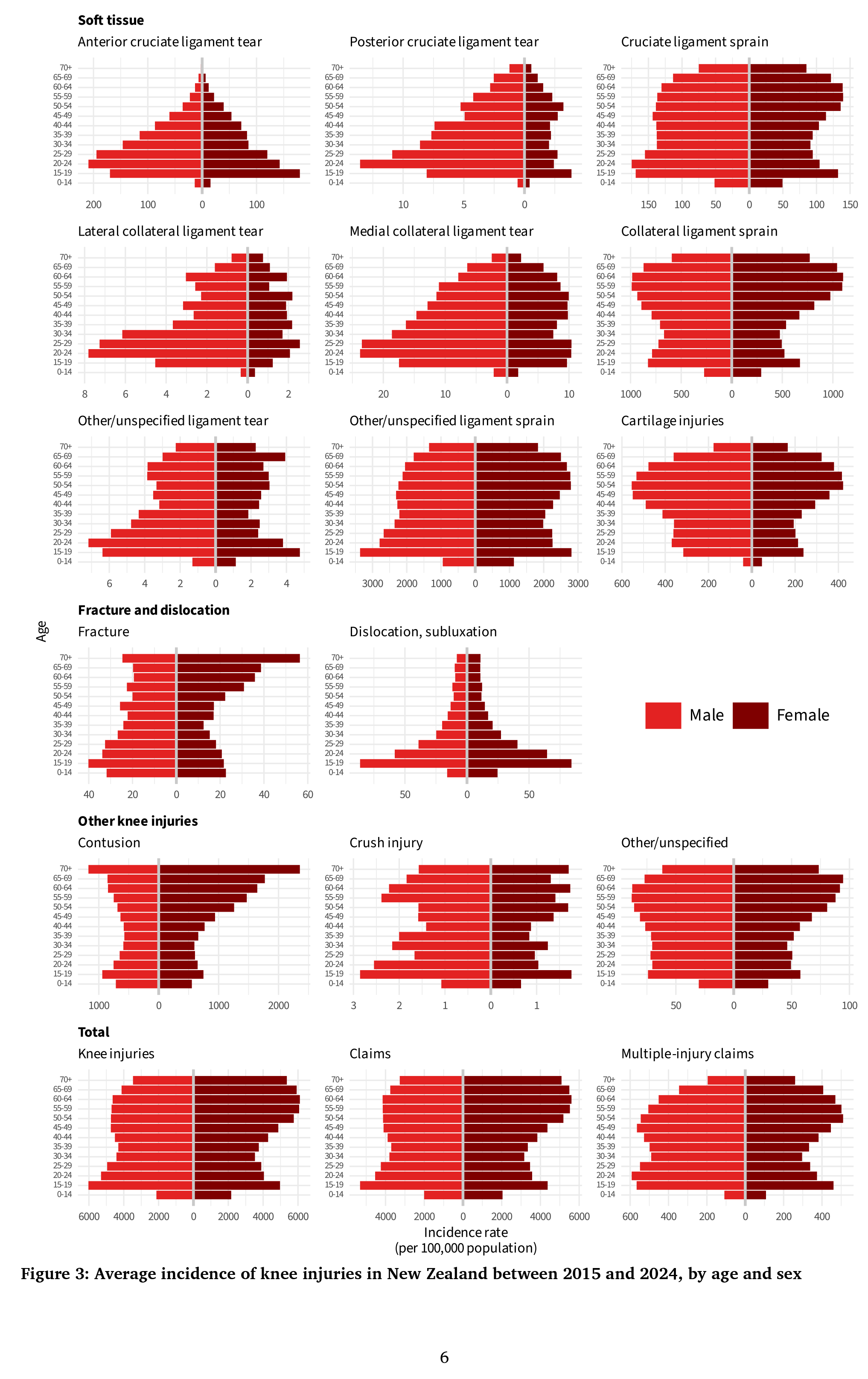
Average incidence of knee injuries in New Zealand between 2015 and 2024, by age and sex

## Discussion and Conclusions

This study provides incidence estimates for a wide range of knee injuries, exploiting comprehensive, whole-of-population data from across all injury settings. Capturing data from primary and community care settings, as well as secondary care and trauma settings, identified a large number of less severe injuries, such as ligament sprains and contusions, that have largely been absent from previous epidemiologic research internationally.

This study is the first to investigate the trends and demographic characteristics of all knee injuries in New Zealand. We found that the total number of knee injuries increased by 32% between 2015 and 2024, while the incidence rate, relative to population size, increased by 9% for males and 22% for females. Females had a higher overall knee injury incidence rate than males (with the gap widening over time), mostly due to higher incidence of other/unspecified ligament sprains and contusions. Lower overall incidence rates for Māori, despite higher rates of ligament tears and dislocations, may be attributable to known barriers to accessing care for Māori^10^, resulting in information bias through missing data from lower severity injuries. Over the life cycle, the highest incidence of injuries was at 15–30 years of age for ligament tears and dislocations, and at 40–65 years of age for cartilage injuries.

The main strength of this study is the use of comprehensive nationwide injury insurance claims records, allowing us to capture most knee injuries on a national level and classify them into major knee injury types. It is, however, important to be aware of potential limitations of the claims dataset, which could be caused, for example, by individuals writing incorrect information on data collection sheets, not making a claim, or not going to a health provider for their injury at all (although there is a high incentive to do so as the scheme provides cover for both medical costs and lost earnings on a “no-fault” basis).

Another limitation is potential misclassification of some knee injuries, due to the large share of injuries coded using ‘other’ or ‘unspecified’ injury codes. Particularly, the numbers of specific ligament injuries may be underestimated, as a large share of ligament injuries were recorded under the category of “other/unspecified ligament sprains”. Soft tissue injuries such as ligament sprains are often treated in primary or community care, where specialised diagnostic equipment (e.g. MRI) may not be available. In this context, providers may choose to use non-specific coding to avoid the possibility of a claim being rejected or treatment coverage being interrupted if a more specific initial diagnosis was later overturned by imaging findings or specialist assessment.

We also note that such misclassification may have increased over time: the ‘other’ or ‘unspecified’ classifications were among the fastest-growing injury types, accounting for 40% of injuries recorded in 2015 but 57% in 2024.

While several previous studies have reported the incidence of knee injuries in different countries and settings, direct comparison with our findings is hampered by differing methodologies and data coverage. Previous evidence from New Zealand reported a rate of 166 to 185 cruciate ligament injuries per 100 000 population between 2007 and 2019^6^, similar to the rates presented here. Peat et al. (2014) used population register data, covering both inpatient and outpatient hospital visits and primary care, to report the incidence of different knee injuries in southern Sweden from 2004 to 20123. The overall rate of injuries was notably lower than in our study, particularly for some injury types such as contusions, cartilage injuries, and collateral ligament sprains. These differences may be due to data coverage (our data includes injuries treated by a range of community and allied healthcare providers including physiotherapists, whereas Peat et al. included primary care doctors and hospital inpatient and outpatient attendance only), inclusion criteria (our data includes all knee injuries recorded, whereas Peat et al. included only one injury per person per year), or breadth of diagnostic coding. Nicholls et al. (2018) reported rates of anterior cruciate ligament tear (confirmed by MRI examination) in Iceland of 75 per 100 000 population^11^, similar to those reported here (66 per 100 000), and similar to the rates for all cruciate ligament sprains and tears reported by Peat et al. (71 per 100 000).

Other previous studies have largely been confined to specific care settings. Gage et al. (2012) reported an incidence of 229 per 100 000 population for knee injuries presenting to emergency departments in the United States between 1999 and 2008^1^. Nordenvall et al. (2012) reported an incidence of 78 per 100 000 for cruciate ligament injuries seen in hospitals (in either in-patient or outpatient settings) in Sweden between 2001 and 2009^7^. Maniar et al. (2022) reported an incidence of (all) knee injuries resulting in hospital admission of 84 per 100 000 and males and 60 per 100 000 for females in Australia in 2017–2018^2^. Despite the differences in coverage, and the corresponding lower overall incidence rates, the demographic distribution of knee injuries in these studies was similar to our findings: males had higher incidence rates of ligament and cartilage injuries than females, while females had higher incidence rate of contusions; the age distributions of incidence rates for different knee injury types were also similar to those reported here. Thus we argue our findings are generalisable internationally, at least to high income, Western countries.

Our findings have several important implications. The numbers and incidence rates of knee injuries are larger than previously reported, implying that the burden on the health system and individuals is larger than previously understood. Our inclusion of comprehensive data from primary and community care settings identified a large number of less severe injuries, such as ligament sprains and contusions, that would likely be missed when relying on hospital-based sources. While these are expected, individually, to have smaller and less long-lasting impacts, in aggregate they represent a substantial burden on patients and the health system.

The large share of non-specific injury coding, accounting for around half of all injuries recorded over the period, is likely due at least in part to the inclusion of data from primary and community care settings, and may be related to concerns about claim acceptance under the ACC scheme. The increase in the number of injuries coded as ‘other’ or ‘unspecified’ injury types (which increased by 87% over the study period, compared to a 5% *decrease* for specifically coded injuries) may be related to changes in claim lodgement over the period, in particular the shift to electronic lodgement and automation of claims acceptance, and suggests a need for renewed emphasis on precise diagnostic coding to inform accurate injury epidemiology.

The breadth of injuries identified here is not fully reflected in current knee injury epidemiological research, which has predominantly focused on cruciate ligament injuries. For example, a large body of research has shown a link between cruciate ligament injuries (4% of recorded injuries in our cohort) and, to a lesser extent, meniscal tears (7%) and the onset of knee osteoarthritis^12^. Other knee injuries, for example collateral ligament injuries (16% of recorded injuries) and contusions (21%), have also been shown to be associated with development of OA^13^, although the evidence base for these is much smaller. Our findings suggest that more research is needed on the incidence, prevention, and short- and long-term outcomes of different knee injuries, beyond a narrow focus on cruciate ligament injuries.

## Data Availability

The individual-level raw data used in the study are not publicly available due to the strict security provisions
of the IDI. Access to the IDI may be made available by Stats NZ to approved researchers. The aggregated
data (by injury/year/sex/ethnicity/age strata), and the code used to construct the tables and figures presented
here, are available via Zenodo (https://doi.org/10.5281/zenodo.22104854).

## Competing Interests

The authors have no competing interests to declare.

## Funding

This research was supported by a grant from the Health Research Council of New Zealand (Programme Grant #22/555).

## IDI Disclaimer

These results are not official statistics. They have been created for research purposes from the Integrated Data Infrastructure (IDI), which is carefully managed by Stats NZ. For more information about the IDI, please visit https://www.stats.govt.nz/integrated-data/.

The results are based in part on tax data supplied by Inland Revenue to Stats NZ under the Tax Administration Act 1994 for statistical purposes. Any discussion of data limitations or weaknesses is in the context of using the IDI for statistical purposes and is not related to the data’s ability to support Inland Revenue’s core operational requirements.

## Appendix A: Supplementary Material

**Table A1:**
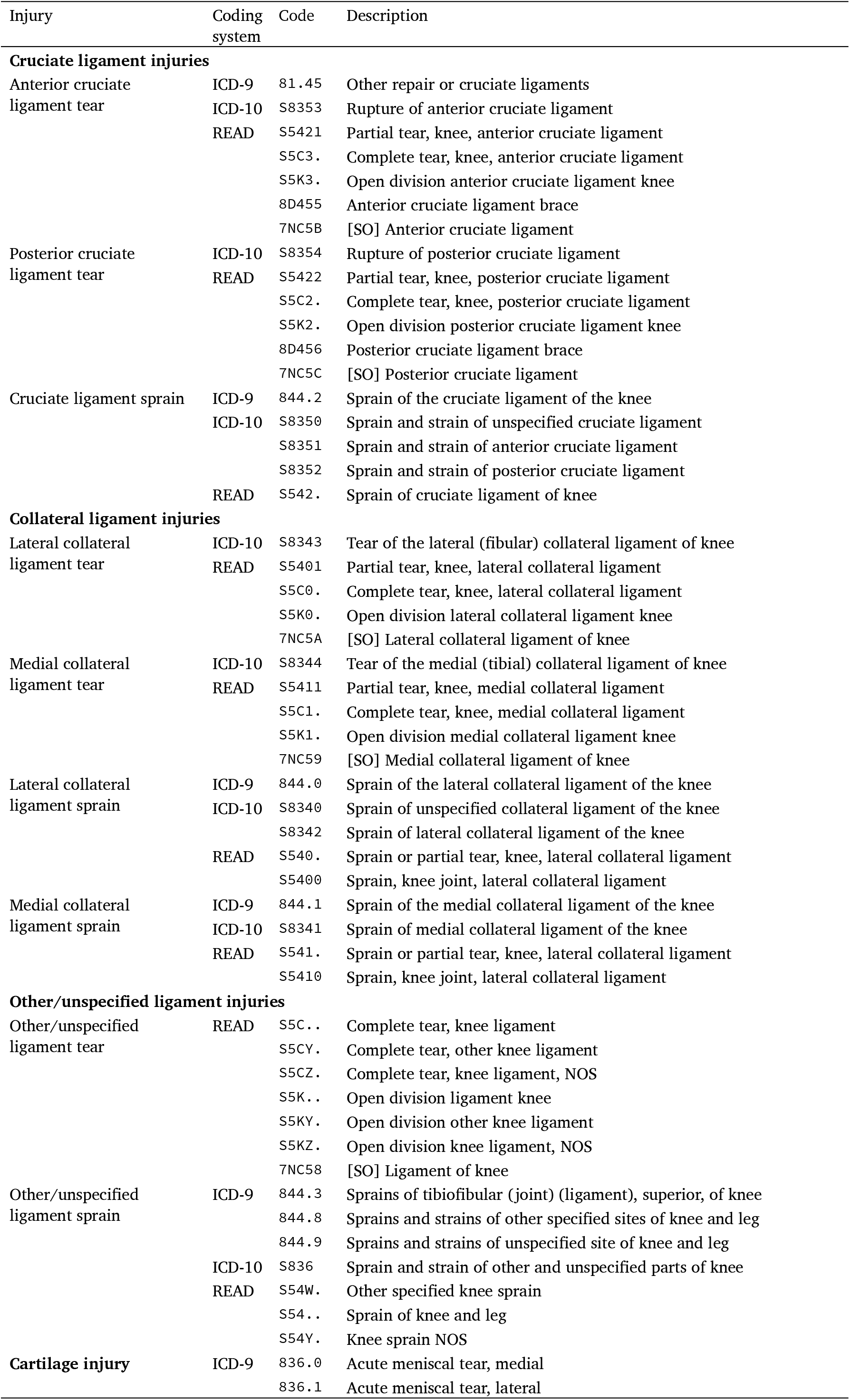
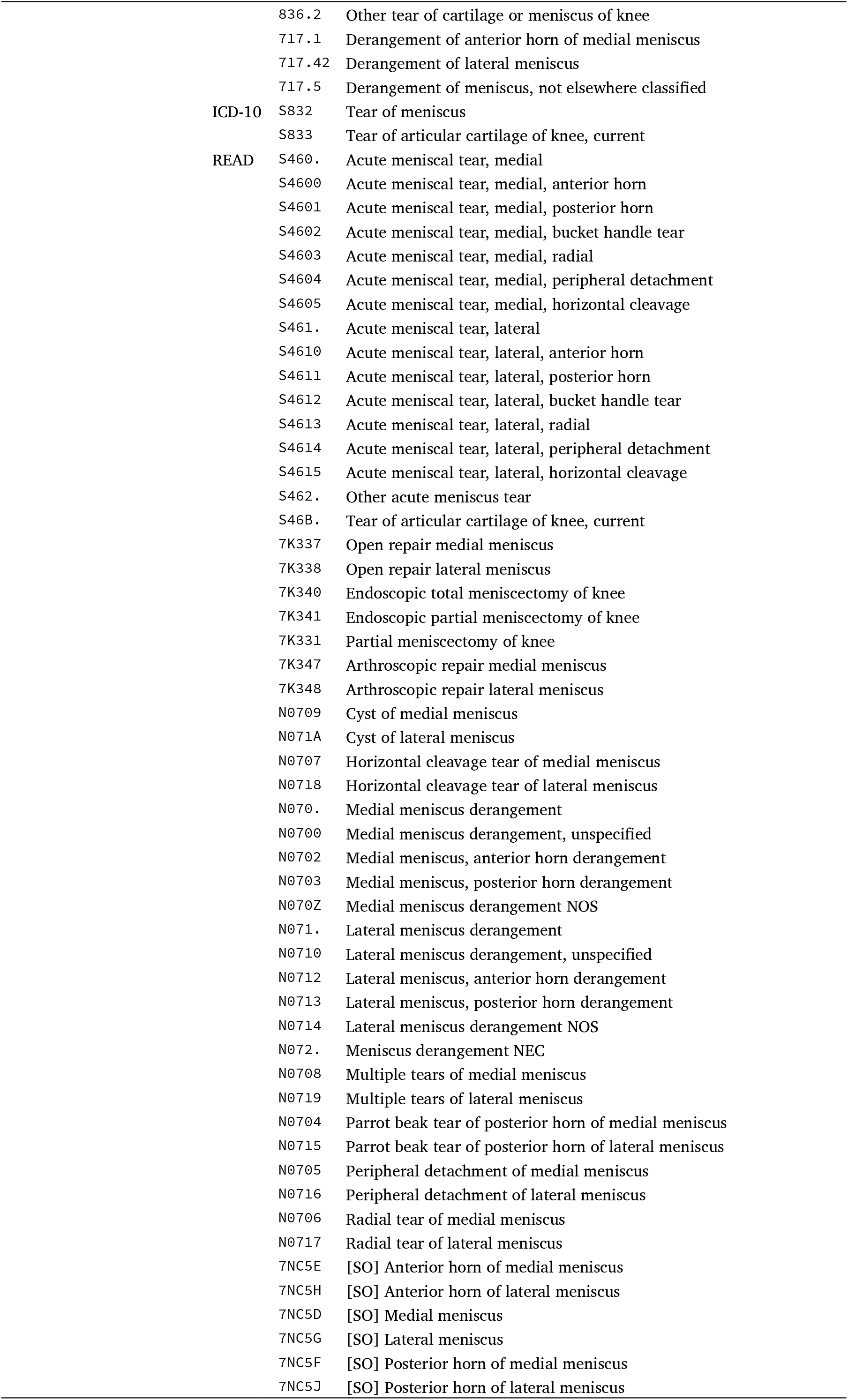
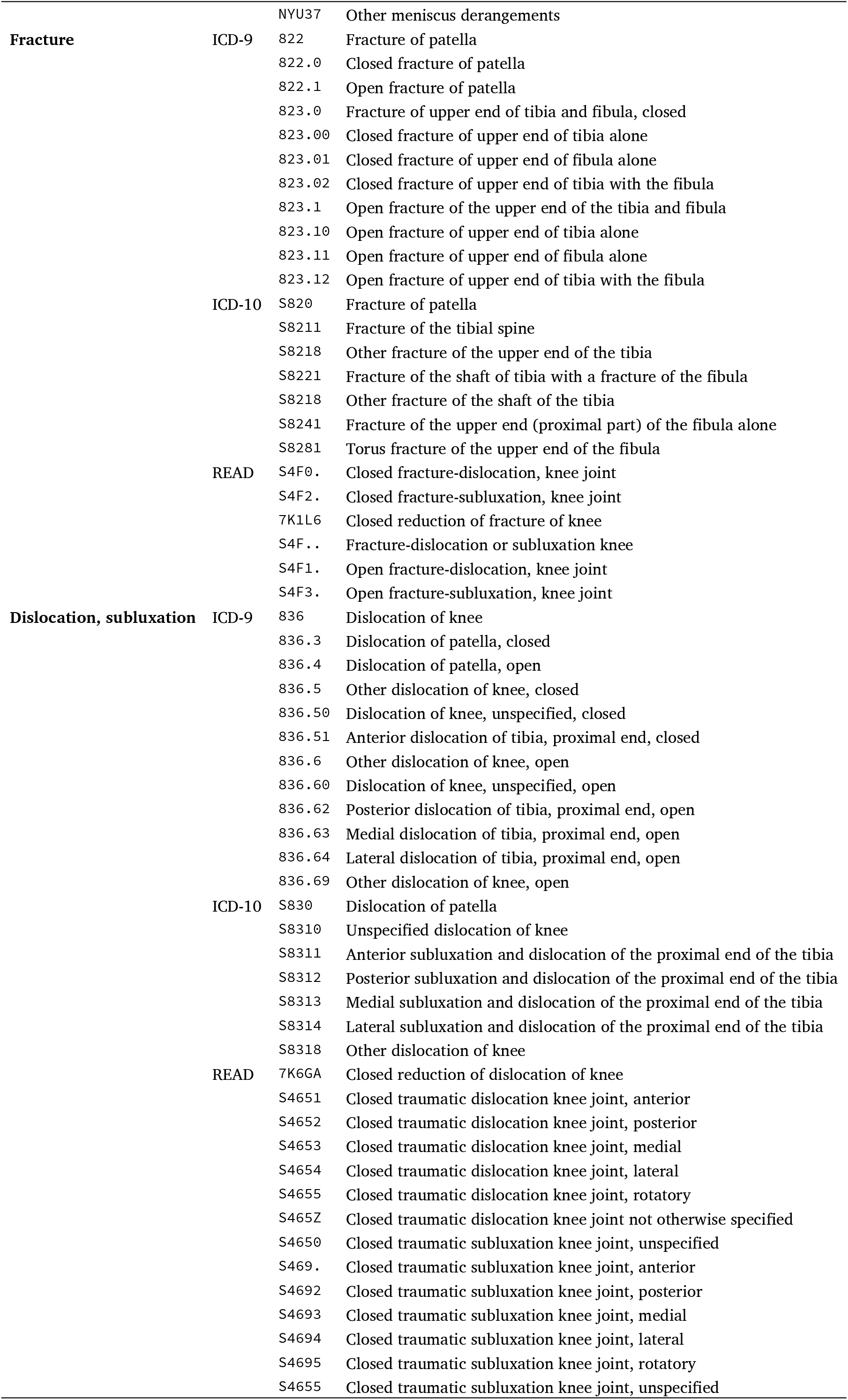
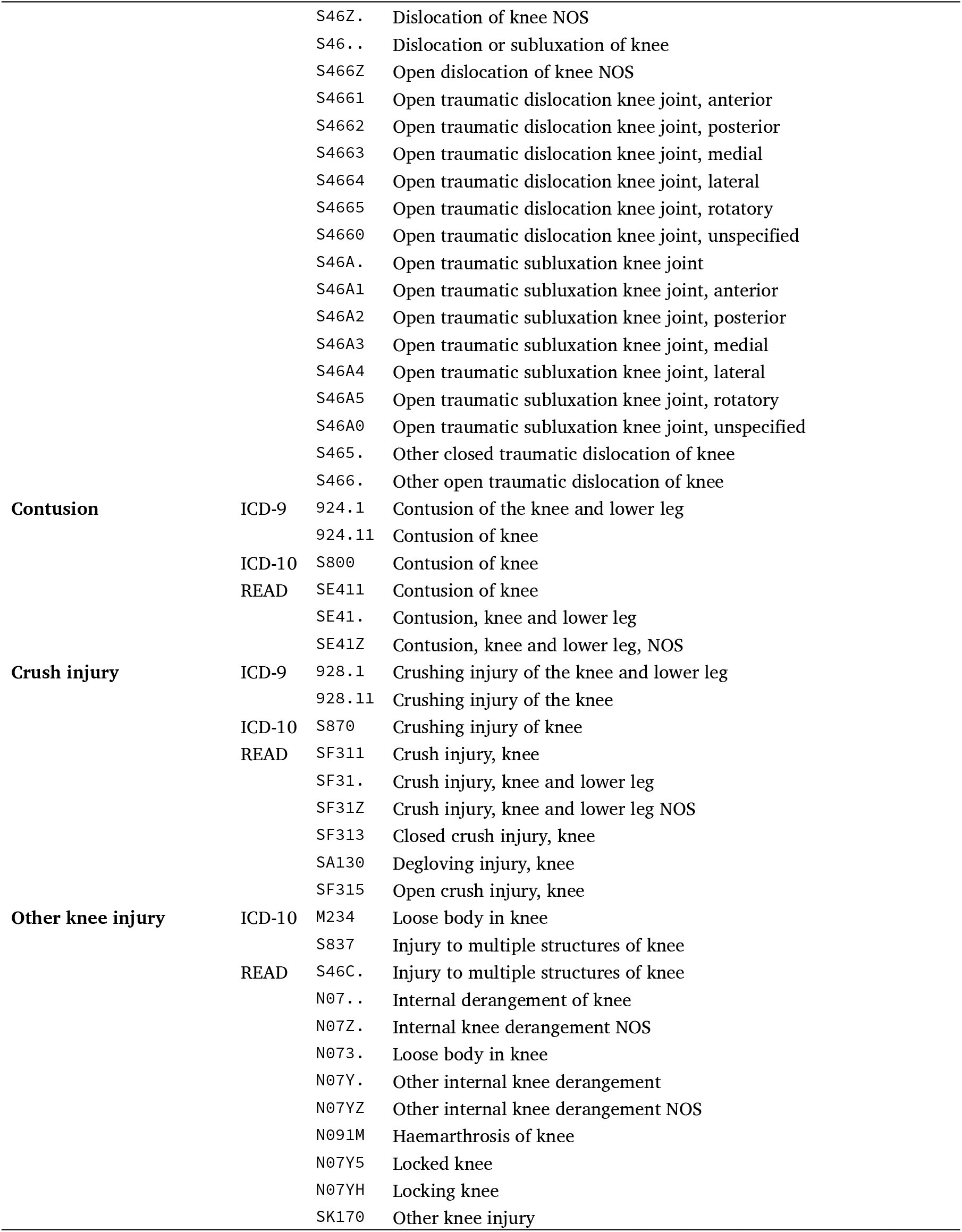
Diagnosis codes used to identify knee injuries.

**Table A2:** Annual incidence of knee injuries in New Zealand per 100,000 population, 2015 to 2024.

| Injury | 2015 | 2016 | 2017 | 2018 | 2019 | 2020 | 2021 | 2022 | 2023 | 2024 |
| --- | --- | --- | --- | --- | --- | --- | --- | --- | --- | --- |
| **Soft tissue injuries** |  |  |  |  |  |  |  |  |  |  |
| *Cruciate ligament injuries* |  |  |  |  |  |  |  |  |  |  |
| ACL tear | 66 | 65 | 65 | 66 | 72 | 59 | 59 | 60 | 69 | 79 |
| PCL tear | 3 | 3 | 3 | 3 | 4 | 3 | 3 | 4 | 4 | 6 |
| Sprain | 136 | 126 | 118 | 120 | 116 | 99 | 103 | 98 | 100 | 103 |
| *Collateral ligament injuries* |  |  |  |  |  |  |  |  |  |  |
| Lateral tear | 3 | 2 | 2 | 2 | 2 | 2 | 2 | 2 | 2 | 2 |
| Medial tear | 8 | 8 | 8 | 8 | 8 | 9 | 9 | 8 | 10 | 13 |
| Sprain | 834 | 819 | 778 | 766 | 738 | 662 | 640 | 588 | 609 | 596 |
| *Other/unspecified ligament injuries* |  |  |  |  |  |  |  |  |  |  |
| Tear | 3 | 3 | 2 | 3 | 2 | 3 | 3 | 4 | 4 | 4 |
| Sprain | 1 589 | 1 719 | 1 783 | 1 853 | 1 973 | 1 905 | 2 001 | 2 124 | 2 356 | 2 627 |
| Cartilage injuries | 346 | 307 | 293 | 300 | 294 | 281 | 282 | 280 | 289 | 260 |
| **Fractures and dislocations** |  |  |  |  |  |  |  |  |  |  |
| Fracture | 34 | 35 | 34 | 33 | 30 | 27 | 24 | 22 | 22 | 20 |
| Dislocation | 28 | 27 | 26 | 26 | 27 | 24 | 25 | 21 | 23 | 24 |
| **Other knee injuries** |  |  |  |  |  |  |  |  |  |  |
| Contusion | 961 | 968 | 959 | 960 | 958 | 857 | 864 | 839 | 892 | 904 |
| Crush injury | 1 | 1 | 1 | 2 | 2 | 1 | 2 | 1 | 2 | 2 |
| Other/unspecified | 55 | 57 | 59 | 61 | 58 | 64 | 68 | 77 | 88 | 61 |
| **Total** | **4 067** | **4 140** | **4 131** | **4 201** | **4 282** | **3 997** | **4 086** | **4 126** | **4 467** | **4 701** |
| Claims | 3 648 | 3 737 | 3 719 | 3 774 | 3 851 | 3 589 | 3 653 | 3 683 | 3 986 | 4 203 |
| Multiple-injury claims | 363 | 350 | 356 | 369 | 370 | 349 | 371 | 379 | 405 | 412 |

Abbreviations: ACL = Anterior cruciate ligament, PCL = Posterior cruciate ligament.

## Notes

### Competing Interest Statement

The authors have declared no competing interest.

### Author Declarations

The Human Research Ethics Committee (Health) of the University of Otago gave ethical approval for this work.

